# Association Between Reduced Handgrip Strength and Commonly Prescribed Medications

**DOI:** 10.1101/2020.01.15.20017087

**Authors:** Bryan L. Love, Erin M. Mitchell, LeAnn B. Norris

## Abstract

Decreased handgrip-strength has become an increasingly important measure of overall health status and fitness. This was a cross-sectional analysis among adult participants in the 2011-2012 NHANES survey. Handgrip-strength was assessed using a digital dynamometer and a standard protocol, and medication use was assessed by self-report and verification by the interviewer. Mean handgrip-strength among participants with no medication use was 87.2kg in males and 57.2kg in females. Handgrip strength decreased significantly in both men and women (ptrend<0.001 for both) with increasing medication use after adjustment for age, sex, height, arm circumference, and BMI. Statins, ACE-inhibitors, ARBs, diuretics, calcium channel blockers, and sulfonylureas showed a consistent and significant decrease in grip strength in linear regression models. In this nationally representative survey of adults in the US, we observed a negative relationship between handgrip strength and polypharmacy. Further, several specific medications, mostly cardiovascular drug classes, were associated with reduced handgrip-strength.

## What is known and Objective

Handgrip strength, or grip strength, is defined as the maximum possible force achieved when hand joints flex due to intrinsic and extrinsic muscles.^1^ It can be measured as either absolute grip strength or relative strength, where the absolute strength is divided by an individual’s Body Mass Index (BMI). Absolute grip strength, also called combined grip strength, is calculated by adding together the best value in both hands. Both combined and relative strength are measured in kilograms (kg) when an individual is standing up.^1, 2^ Decreased grip strength relative to others of the same age and sex is correlated with increased risk of death from all causes.^3-6^ In addition, poor grip strength correlates with an increased risk of hospitalization and disability.^7^ People with low grip strength relative to their peers may meet the clinical definition of sarcopenia, or be at risk of developing it as they age.^8^ Sarcopenia is a gradual loss of strength and skeletal muscle; loss of muscle without loss of strength or physical function may be considered pre-sacropenia.^8, 9^

Age-related declines in skeletal muscle mass, strength, and quality is known as sarcopenia. The extent that medications induce or worsen sarcopenia is unclear, but it is important to physicians and other healthcare providers Certain medications are taken more frequently in those with lower grip strength values; thus, there is considerable interest in the role that medications may play in grip strength.^10^ Polypharmacy, defined in many studies as the use of four or more daily medications, has been associated with reduced grip strength in some studies, although it is not known if specific medications may have played a greater role than quantity alone.^10, 11^ The pharmacology of several medication classes can impact intracellular processes that affect muscle function. For example, calcium channel blockers slow calcium influx, diuretics cause excretion of intracellular cations (Na^+^, K^+^, Ca^++^), and statins cause muscle cramping as a common side effect.^12, 13^ The purpose of the present study was to evaluate the association between commonly prescribed medications with known or hypothesized skeletal muscle effects and combined grip strength using data from the NHANES study.

## Methods

### Study design and participants

This study involved a cross-sectional analysis of adults who participated in the examination component of NHANES, a complex, multistage, probabilistic survey conducted by the Centers for Disease Control and Prevention and the National Center for Health Statistics. We examined NHANES data from 2011 to 2012 and we limited our analysis to participants aged 30 years or older, due to the higher prevalence of chronic medication use in this population.

### Compliance with Ethical Standards

The NHANES survey is approved by the institutional review board of the National Center for Health Statistics. Oral and written informed consent was obtained from all participants. All authors report no relevant conflicts of interest.

### Grip strength

Grip strength was assessed using a standardized protocol following a demonstration by a trained examiner. A digital grip strength dynamometer (Takei Model # T.K.K.5401) was used for all assessments. Subjects were instructed to stand, unless physically unable, and hold the dynamometer parallel to their side. Each participant was randomly assigned to start the test with their dominant or non-dominant hand. Following a demonstration and practice, the participant was asked to use one of their hands to squeeze the dynamometer as hard as possible while exhaling. The participant alternated between each hand with a 1-minute rest between measurements on the same hand with a goal of three recorded measurements for each hand. The combined grip strength was recorded as the sum of the best measurement from the right and left hand. Subjects who were only able to complete the assessment with one hand were excluded from the analysis. A detailed grip strength procedure manual is available for download at the NHANES website.^14^

### Medications

NHANES participants were asked questions about prescription medication use by trained interviewers using a computer assisted interview program. During the interview, survey participants were asked: “have you taken any medications in the past 30 days for which they needed a prescription.” If participants answered “yes”, they were asked to show the interviewer the medication containers for all the medications taken. The medication name was recorded, and the participant was asked how long they had taken the medication. The total number of prescription medications was also recorded and used to create a dichotomous “polypharmacy” variable, defined as taking five or more medications each day. Medications of interest were queried by searching for generic name or the NHANES-assigned drug id. A binary variable was created for each medication or medication class.

### Statistics

We generated nationally representative estimates after incorporating appropriate 2-year sample weights, which accounts for the sampling design of NHANES. Robust linearized standard errors and 95% confidence intervals were obtained using the “svy” prefix with “subpop” option for subpopulations, and the “over” option for between-group comparisons (such as females vs. males) using Stata/MP 15.1 (StataCorp, College Station, TX). The 2-sided statistical significance level was set at α=0.05.

Sociodemographic characteristics, body measurements, and the prevalence of medication use in the sample population were presented as weighted mean or weighted percentages ± standard error for continuous and categorical variables, respectively. Age, sex, arm circumference, arm length, physical activity, smoking status, and BMI were evaluated as potential covariates to include in the adjusted multivariable models. We started with an empty model and used a pragmatic forward selection process adding or dropping an individual variable and re-estimating the model at each step. Once the baseline linear regression model was selected, we then evaluated the effect of each medication variable. To assess the association between medication classes and combined grip strength, simple and multivariable linear regression models were conducted. A similar approach was used to examine the effect of polypharmacy, and we used the same covariates in a multivariable model with a variable measuring the count of current medications. Average values for combined grip strength were calculated for men and women separately using the polypharmacy model at fixed values of the covariates. P-values for trend were calculated from these adjusted linear regression models.

## Results

### Summary characteristics

Weighted characteristics of the 2,230 males and 2,336 females are presented in Table 1. Age and most physical characteristics were significantly different between males and females as expected. Combined grip strength was significantly different between males and females (87.98 vs. 55.2 kg; p<0.001). There were fewer current smokers among females, and they were less likely to have ever smoked compared to male participants. A composite variable for heart disease was more common in males, although this was not statistically significant. Coronary heart disease (4.77% vs. 2.54%); p<0.001) and myocardial infarction (4.72% vs. 3.07%; p=0.02) were more common in males compared to females, respectively. Heart failure, stroke, and angina pectoris were similar between males and females. There was no significant difference in physical activity, BMI, or the prevalence of diabetes between men and women.

**Table 1:** Weighted Characteristics of Adults > 30 years of Age by Sex^1^.

|  | Men | Women | p-value |
| --- | --- | --- | --- |
| N | 2,230 | 2,336 |  |
| Age, yrs | 52.1 (0.64) | 53.2 (0.45) | 0.02 |
| Combined grip strength, kg | 88.4 (0.93) | 55.5 (0.46) | <0.001 |
| Height, cm | 175.5 (0.34) | 161.7 (0.36) | <0.001 |
| Arm circumference, cm | 34.3 (0.13) | 32.3 (0.19) | <0.001 |
| Upper arm length, cm | 39.1 (0.10) | 35.9 (0.11) | <0.001 |
| BMI, kg/m<sup>2</sup> | 28.9 (0.22) | 29.3 (0.27) | 0.11 |
| Physical activity<sup>2</sup>, % | 87.8 (1.61) | 86.4 (1.64) | 0.62 |
| Ethnicity, % |  |  |  |
| Mexican American | 7.46 (1.59) | 6.38 (1.49) |  |
| Other Hispanic | 5.81 (1.39) | 6.17 (1.67) | 0.18 |
| Non-Hispanic White | 69.25 (3.53) | 68.21 (3.77) |  |
| Non-Hispanic Black | 10.18 (2.06) | 11.90 (2.37) |  |
| Other race, including multi-racial | 7.29 (1.03) | 7.34 (0.99) |  |
| Diabetes<sup>3</sup>, % | 11.46 (0.78) | 10.59 (0.81) | 0.35 |
| Heart disease<sup>4</sup>, % | 11.15 (0.80) | 9.49 (0.50) | 0.06 |
| Smoking status, % |  |  |  |
| Current smoker | 23.1 (1.48) | 15.6 (1.26) |  |
| Former smoker | 30.5 (1.72) | 24.2 (1.93) | <0.001 |
| Never smoker | 46.4 (1.97) | 60.3 (1.39) |  |
| Prescription medications, # | 2.08 (0.13) | 2.46 (0.10) | <0.01 |
| Met criteria for polypharmacy, % | 16.0 (1.58) | 19.5 (1.23) | 0.06 |
| Prescription medication use, % |  |  |  |
| HMG-CoA reductase inhibitors (i.e. Statins) | 21.7 (1.24) | 20.4 (1.40) | 0.39 |
| ACE inhibitors | 16.3 (0.92) | 12.4 (0.79) | <0.001 |
| Diuretics | 12.1 (1.02) | 17.51 (0.91) | <0.001 |
| Beta blockers | 12.2 (1.37) | 14.1 (1.10) | 0.22 |
| Hypothyroid medications | 3.9 (0.66) | 10.9 (0.91) | <0.001 |
| Calcium channel blockers | 8.4 (0.88) | 7.6 (0.92) | 0.50 |
| Angiotensin receptor blockers | 5.9 (0.74) | 8.2 (0.69) | 0.03 |
| NSAIDs | 3.9 (0.57) | 7.0 (0.85) | <0.01 |
| Sulfonylureas | 4.6 (0.46) | 3.3 (0.73) | 0.08 |
| Fibrates | 2.2 (0.59) | 1.1 (0.19) | 0.02 |
| Xanthine oxidase inhibitors (e.g., Allopurinol) | 1.78 (0.48) | 0.44 (0.16) | 0.02 |
| Antiarrhythmics | 1.2 (0.29) | 1.2 (0.23) | 0.99 |
| Nitrates | 0.54 (0.20) | 0.73 (0.12) | 0.47 |

1. Characteristics of participants are given as percentage (standard error) in each category (e.g. diabetes) or weighted arithmetic mean (standard error) in each continuous variable (e.g. age).
2. Dichotomous variable indicating recreational activity of at least 150 minutes per week of moderate intensity or 75 minutes per week of vigorous intensity.
3. Survey respondents were asked “Other than during pregnancy, have you ever been told by a doctor or health professional that you have diabetes or sugar diabetes?”
4. Combined variable from respondents who answered the following questions: “Has a doctor or other health professional ever told you that you had {coronary heart disease, angina/angina pectoris, congestive heart failure, heart attack, stroke}?”

### Prescription medication use

Overall, 43.4% of men and 54.9% of women reported taking at least one prescription medication, and 80.1% of participants’ medication containers were viewed by the interviewer. Men reported an average of 2.08 medications compared to 2.46 among women (p=0.002), but there was no statistical difference between men and women in the proportion of participants who met the criteria for polypharmacy (16.0% vs. 19.5%; p=0.058). The top four most commonly reported medications included HMG-CoA reductase inhibitors, ACE inhibitors, diuretics, and beta-blockers.

### Effect of medications on grip strength

Physical activity, smoking status, and arm length were evaluated as potential covariates, but these were excluded due to lack of statistical significance in the baseline multivariable model. After adjustment for age, sex, height, arm circumference, and BMI, grip strength decreased significantly as the number of prescription medications increased in both men and women (Figure 1). In men, the mean combined grip strength was 87.2 kg (95% CI 85.3-89.0) without any medication use but decreased to 80.2 kg (95% CI 76.9-83.6) with 10 or more medications. Similar effects were noted among females with mean combined grip strength decreasing from 57.2 kg (95% CI 56.0-58.4) with no medication use to 50.3 (95% CI 47.3-53.3) with 10 or more medications.

**Figure 1:**
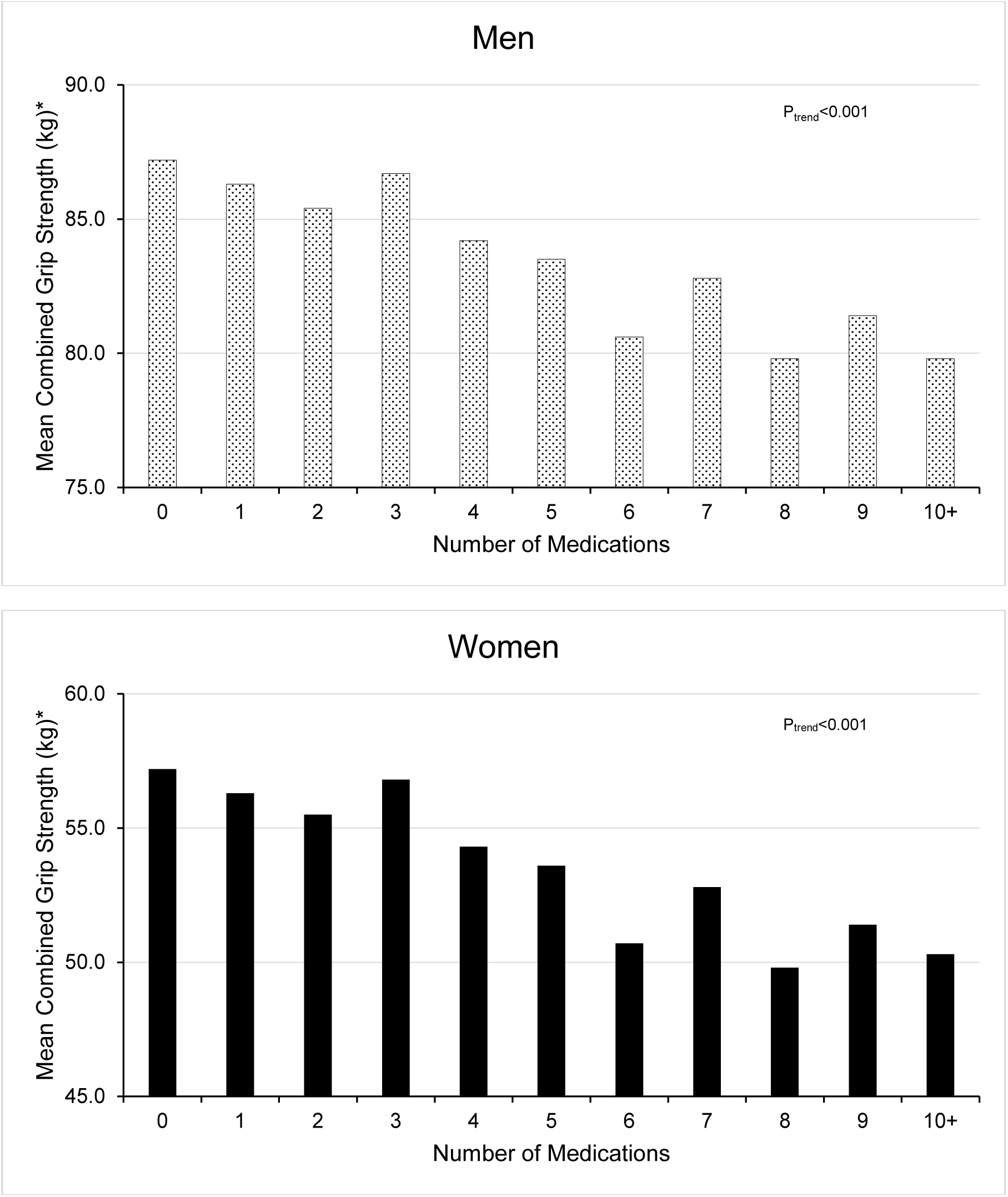
Average combined grip strength according to number of medications reported and stratified by gender. *Adjusted for age, height, BMI, and arm circumference. P values for trend are from adjusted linear regression models.

In bivariate linear regression analyses, all medication classes studied demonstrated significant negative associations with combined grip strength with the exception of fibrates and allopurinol (Table 2). The overall effect on grip strength was lessened for all medication classes in multivariable linear regression models adjusting for age, sex, height, arm circumference and BMI. Statins, ACE-inhibitors, ARBs, diuretics, calcium channel blockers, and sulfonylureas showed a consistent and significant decrease in grip strength in both unadjusted and multivariable linear regression models. This effect was most prominent in reported users of sulfonylurea medications (-4.80 kg) followed by users of angiotensin receptor blockers (-3.85 kg). Within the class of diuretics, users of loop diuretics (e.g. furosemide) had the largest significant decrease in grip strength (-5.37 kg; p<0.001), and users of thiazide (-0.98 kg) and potassium-sparing (-1.17 kg) diuretics had decreases in grip strength that were not statistically significant in multivariable models.

**Table 2:** Linear Regression Models of Grip Strength and Medication Use.

|  | Model 1<sup>a</sup> |  | Model 2<sup>b</sup> |  |
| --- | --- | --- | --- | --- |
|  | Coefficient | p-value | Coefficient | p-value |
| HMG-CoA reductase inhibitors | -8.47 | <0.001 | -1.99 | 0.03 |
| ACE inhibitors | -2.70 | 0.04 | -1.61 | 0.03 |
| Angiotensin receptor blockers | -13.45 | <0.001 | -3.85 | <0.01 |
| Beta-blockers | -8.47 | <0.001 | -0.93 | 0.30 |
| Diuretics | -9.21 | <0.001 | -1.91 | 0.05 |
| Calcium channel blockers | -8.06 | <0.001 | -1.99 | <0.01 |
| Nitrates | -14.94 | 0.05 | -4.39 | 0.13 |
| Fibrates | -1.15 | 0.97 | -1.59 | 0.49 |
| NSAIDs | -6.13 | <0.01 | -1.41 | 0.07 |
| Antiarrhythmics | -16.27 | <0.01 | -4.01 | 0.08 |
| Hypothyroid medications | -13.30 | <0.001 | -0.90 | 0.55 |
| Hyperthyroid medications | -19.21 | 0.02 | 2.46 | 0.15 |
| Sulfonylureas | -9.20 | <0.001 | -4.80 | 0.03 |
| Xanthine oxidase inhibitors | 3.07 | 0.406 | -2.16 | 0.26 |

<sup>a</sup> Unadjusted, simple linear regression
<sup>b</sup> Adjusted for covariates (age, sex, height, arm circumference, arm length, and BMI).

## Discussion

The grip strength results from the current study of NHANES 2011-2012 participants share confirmatory and conflicting information with those of some other studies. The KORA-Age study, a study that tracked 711 persons with at least two chronic health conditions, did not find a statistically significant correlation between polypharmacy (>4 medications) and reduced grip strength. However, they did not evaluate medication classes in isolation; other studies have found relationships between grip strength and specific medications.^15^ Anti-hypertensive medications are most commonly associated with low grip strength. The Hertfordshire Cohort Study (HCS) showed that furosemide and nitrates were associated with reduced grip strength, but did not find any significant relationship between angiotensin converting enzyme (ACE) inhibitors, thiazides, or statins and decreased grip strength.^16, 17^ A second study evaluated anti-adrenergic medications and ACE inhibitors using data from the 1999-2002 and 2011-2012 National Health and Nutrition Examination Survey (NHANES) and found that those who took central- and peripherally-acting antiadrenergic agents had decreased combined grip strength.^18^ However, other anti-hypertensives such as beta blockers, calcium channel blockers, and diuretics were not evaluated. Although few studies have focused specifically on grip strength, statins are known to affect muscle function and inhibit physical activity -- a commonly cited reason for discontinuation, especially among elderly.^19^ Both the present study using NHANES data and the HCS study found that loop diuretics significantly decreased grip strength. Our findings also suggest that statins and ACE-inhibitors are also associated with decreased grip strength. In contrast, HCS did not find a statistically significant relationship between statins and ACE-inhibitors and decreased grip strength, and only found a relationship between calcium channel blockers and grip strength in women.^18^ However, HCS’s total study population was smaller (HCS: 2,987 vs. NHANES: 4,566) and HCS adjusted physical activity level of participants.^16^ Physical activity was evaluated as a covariate in our study, but was not a significant predictor of combined grip strength in the baseline model selection process.

Multiple factors appear to be at work in medication-induced sarcopenia, including an increase in inflammation and a decrease in production of androgens and growth hormone, both of which promote skeletal muscle growth. Aging influences this to some extent, but medications which decrease grip strength may alter these processes by increasing the activity of pro-inflammatory cytokines. These cytokines promote muscle atrophy by causing myoproteins to degrade faster than they can be rebuilt.^9^ As a result, the number of muscle fibers and motor units decrease, causing a decrease in muscle mass and strength that may lead to sarcopenia over time.^8^ In contrast, mouse studies have suggested that a decrease in satellite cells (stem cells of skeletal muscle) reduces the regenerative capacity of myoprotein but does not directly correlate with a reduction in strength or function as the aging process itself does.^20^ However, it is important to note that certain diseases, including obesity and cardiovascular disease for which medications like statins and anti-hypertensives might be prescribed, may increase inflammation and could cause muscle wasting even without the use of medications.^9^

Since five out of the six medications that showed a consistent decrease in grip strength target cardiovascular health conditions, prescribing providers need to weigh the potential benefit of these medications carefully against adverse effects, including decreased grip strength and other troubling side effects. However, since grip strength decrease was shown for those taking 10 or more medications, this “polypharmacy effect” highlights a need for providers to monitor patients’ complete medication list and weigh benefits vs. risks.

Our study has several limitations. Since NHANES data are cross-sectional, causality cannot be implied as there are not longitudinal assessments of grip strength pre- and post-medication initiation. Secondly, we have attempted to control for several factors associated with grip strength; however, residual confounding may still be present. It is difficult to disentangle the direct effects of medications and chronic diseases on muscle which may negatively impact grip strength. Finally, we conducted several statistical tests to assess the effect of individual medications on grip strength which creates the possibility of a type I error or false positive results by chance alone. However, this seems unlikely since our findings were similar to previous reports.

Despite these limitations, our study had several strengths. We analyzed a large database of non-institutionalized men and women that is nationally representative of the US population. The data were collected according to a standardized protocol. Medications were assessed by trained interviewers and pill bottles were verified in most cases. Grip strength was assessed using strict protocols and quality control.

## What is new and Conclusion

In summary, we observed a negative relationship between grip strength and medication use. This effect was noted with increasing quantity of medication use (i.e. polypharmacy) as well as with specific medication classes. These effects may reflect the effect of underlying diseases or direct effects of medications on muscle mass and function. Our findings confirm prior observational studies, and suggest that additional research is needed to determine if a causal relationship exists.

## Data Availability

All data utilized in the analysis is freely available at the NHANES website.

https://wwwn.cdc.gov/nchs/nhanes/default.aspx

## References

1. Norman K, Stobäus N, Gonzalez MC, Schulzke JD, Pirlich M. Hand grip strength: outcome predictor and marker of nutritional status. Clin Nutr 2011;2:135–42.

2. Lawman HG, Troiano RP, Perna FM, Wang CY, Fryar CD, Ogden CL. Associations of Relative Handgrip Strength and Cardiovascular Disease Biomarkers in U.S. Adults, 2011-2012. Am J Prev Med 2016;6:677–83.

3. Leong DP, Teo KK, Rangarajan S, et al. Prognostic value of grip strength: findings from the Prospective Urban Rural Epidemiology (PURE) study. Lancet 2015;9990:266–73.

4. Volaklis KA, Halle M, Meisinger C. Muscular strength as a strong predictor of mortality: A narrative review. Eur J Intern Med 2015;5:303–10.

5. Volaklis KA, Halle M, Koenig W, et al. Association between muscular strength and inflammatory markers among elderly persons with cardiac disease: results from the KORA-Age study. Clin Res Cardiol 2015;11:982–9.

6. Gale CR, Martyn CN, Cooper C, Sayer AA. Grip strength, body composition, and mortality. Int J Epidemiol 2007;1:228–35.

7. Legrand D, Vaes B, Matheï C, Adriaensen W, Van Pottelbergh G, Degryse JM. Muscle strength and physical performance as predictors of mortality, hospitalization, and disability in the oldest old. J Am Geriatr Soc 2014;6:1030–8.

8. Burton LA, Sumukadas D. Optimal management of sarcopenia. Clin Interv Aging 2010;217–28.

9. Campins L, Camps M, Riera A, Pleguezuelos E, Yebenes JC, Serra-Prat M. Oral Drugs Related with Muscle Wasting and Sarcopenia. A Review. Pharmacology 2017;1-2:1–8.

10. Jensen LD, Andersen O, Hallin M, Petersen J. Potentially inappropriate medication related to weakness in older acute medical patients. Int J Clin Pharm 2014;3:570–80.

11. König M, Spira D, Demuth I, Steinhagen-Thiessen E, Norman K. Polypharmacy as a Risk Factor for Clinically Relevant Sarcopenia: Results From the Berlin Aging Study II. J Gerontol A Biol Sci Med Sci 2017;1:117–22.

12. Shields DL. Calcium channel blockers as initial therapeutic agents in hypertension: relationship to incident heart failure. Biol Res Nurs 2014;3:266–77.

13. Taylor BA, Thompson PD. Muscle-related side-effects of statins: from mechanisms to evidence-based solutions. Curr Opin Lipidol 2015;3:221–7.

14. (NHANES) NHaNES. Muscle Strength Procedures Manual, Available from https://wwwn.cdc.gov/nchs/data/nhanes/2011-2012/manuals/muscle_strength_proc_manual.pdf. Accessed February 2, 2018.

15. Volaklis KA, Thorand B, Peters A, et al. Physical activity, muscular strength, and polypharmacy among older multimorbid persons: Results from the KORA-Age study. Scand J Med Sci Sports 2018;2:604–12.

16. Ashfield TA, Syddall HE, Martin HJ, Dennison EM, Cooper C, Aihie Sayer A. Grip strength and cardiovascular drug use in older people: findings from the Hertfordshire Cohort Study. Age Ageing 2010;2:185–91.

17. Witham MD, Syddall HE, Dennison E, Cooper C, McMurdo ME, Sayer AA. ACE inhibitors, statins and thiazides: no association with change in grip strength among community dwelling older men and women from the Hertfordshire Cohort Study. Age Ageing 2014;5:661–6.

18. Loprinzi PD, Loenneke JP. The effects of antihypertensive medications on physical function. Prev Med Rep 2016;264–9.

19. Banach M, Serban MC. Discussion around statin discontinuation in older adults and patients with wasting diseases. J Cachexia Sarcopenia Muscle 2016;4:396–9.

20. Fry CS, Lee JD, Mula J, et al. Inducible depletion of satellite cells in adult, sedentary mice impairs muscle regenerative capacity without affecting sarcopenia. Nat Med 2015;1:76–80.

